# PROgressive struCturEd Simulation-based Surgical training program (PROCESS) - Open Vascular Surgery: Study protocol for triple-arm, randomized, single-blinded educational clinical trial

**DOI:** 10.1101/2024.08.22.24312415

**Authors:** Alejandro Velandia-Sánchez, Camilo A. Polanía-Sandoval, José V. Álvarez-Martínez, Santiago Uribe-Ramírez, Juliana Tello-Pirateque, Carlos J. Pérez-Rivera, Juan P. Ávila-Madrigal, Danna L. Cruz Reyes, Paulo A. Cabrera-Rivera, Camilo E. Pérez-Cualtan, Edgar C. Barrera, Yury F. Bustos-Martínez, Sebastián Gómez Galán, Juan C. Briceño, Michel M.P.J. Reijnen, Jaime Camacho-Mackenzie, Carlos O. Mendivil, Juan G. Barrera-Carvajal

## Abstract

**Introduction:** Vascular surgery has been directed towards endovascular approaches; however, not all patients qualify for these procedures. Open vascular surgery remains crucial, demanding a steep learning curve. Exposure to these procedures has declined, resulting in a need for more standardization in acquiring open vascular surgery skills and potentially contributing to poorer outcomes. Simulation offers a solution, yet the evidence for structured programs in open vascular surgery is limited. This study aims to compare the efficacy of technical skill acquisition between a structured, progressive simulation-based training program and traditional experience-based training in open vascular surgery.

**Methods:** A randomized, single-blinded, triple-arm educational clinical trial will be conducted. A control and intervention phases of three groups with different exposure levels to the simulation program are proposed. Group 1: open abdominal aortic repair, Group 2: vascular anastomosis and open abdominal aortic repair, and Group 3: specific surgical skills, vascular anastomosis, and open abdominal aortic repair. The 3D-printed models from AngioCT will be used for the open abdominal aortic repair simulation. Surgical residents of general, vascular, or cardiothoracic surgery programs will be included. Sample size calculation resulted in 45 participants, 15 per group. Single blinding will involve external evaluators. Randomization will occur as a stratified randomization.

**Discussion:** We expect that the structured and progressive simulation-based training program would enhance technical surgical skills. Based on the progression through different modules within the program, we aim to evaluate differences in the acquisition of technical surgical skills. We hypothesize that 3D-printed patient-specific models can enhance participants’ vascular surgery training and provide optimal simulated scenarios while prioritizing patient safety. We hope this initiative will impact the formation of future vascular surgeons, shape future training programs, and ensure comprehensive preparation for open vascular surgery.

**Trial registration:** This study protocol was registered in clinicaltrials.gov with the NCT-ID: NCT06452901.

## Introduction

Technological and scientific advancements in vascular surgical procedures have significantly shifted toward endovascular surgery (1). The most notable case is the management of Abdominal Aortic Aneurysms (AAA), where Endovascular Aneurysm Repair (EVAR) has revolutionized treatment by avoiding aortic clamping, reducing bleeding rates, shortening lower limb ischemia time, and minimizing changes in cardiac indexes and silent ischemic events (2,3). These advantages result in lower in-hospital mortality, shorter hospital stays, and faster recovery (4). Consequently, the use of EVAR for unruptured AAAs increased from 45% in 2004 to 83% in 2015 in the USA (5). However, not all patients are candidates for EVAR, resulting in the continued use of Abdominal Aortic Aneurysm Open Repair (AAOR) as the preferred treatment option in such cases (1,4).

Performing an AAOR requires effective teamwork and a vascular surgeon with highly specific surgical skills (6). There is an inversely proportional relationship between the mortality of this procedure and the volume of patients managed by the institution and the surgeon (7). Interestingly, the mortality rate for AAOR increased from 3.2% to 4.1% between 2005-2009 and 2010-2013 (p = 0.008)(6).

This increase in mortality rates can be partially attributed to the decreasing exposure to open vascular surgery (OVS) due to several factors, including the adoption of endovascular techniques, reduced working hours, and improved patient safety protocols (7). Globally, these factors have been noted to hinder the acquisition of technical surgical skills in OVS through experience-based learning (8). Consequently, the lack of standardized training and competencies due to inequitable opportunities to assist in OVS could be one of the multiple factors affecting surgical outcomes in AAOR (8).

To address this issue, simulation-based training has emerged as a crucial tool for acquiring surgical technical skills, positioning itself as essential in training various surgical specialties (9,10). The traditional educational model of “*See one, Do one, Teach one*” is being replaced by “*Do many on a simulator, attain competency, then perform under supervision in the operating room*”(8).

In OVS, only isolated interventions such as simulations of vascular anastomoses (VA), AAOR, or carotid endarterectomies have been implemented, showing high efficacy in acquiring technical surgical skills (7,11,12). However, the evidence primarily consists of heterogeneous observational studies conducted by North American and European trainees (7). Despite their validity, these methodologies are based on individual learning structures and do not encompass comprehensive training programs (10). This shows the need for a structured, progressive, simulation-based program in OVS.

## Methods/Design

### Aims of the study

The primary objective is to compare the efficacy of a progressive, structured simulation-based training program versus traditional experience-based training in acquiring technical surgical skills in OVS among residents in general, vascular, and cardiothoracic surgery.

Secondary objectives include designing and implementing a micro-curriculum for progressive structured simulation-based training in OVS; assessing technical surgical skills in OVS before and after the implementation of the structured training program; analyzing differences in skill acquisition attributed to the varying levels of simulation exposure among different groups within the program; and evaluating the reaction and satisfaction of residents with the structured, progressive simulation-based training program in OVS.

### Study setting

The study will be conducted at the advanced simulation center of the Universidad del Rosario in Bogotá,

DC., Colombia. Participants will include residents from any year of general, vascular, and cardiothoracic surgical programs from various institutions in Bogotá.

### Study design

An experimental, three-arm, randomized, single-blinded clinical trial will be conducted. The SPIRIT guidelines were followed (**Fig 1 and S1 Appendix)**(13). Due to its educational nature, a control and intervention phase will be conducted in all three arms (**Figure 2**). This allows all participants to have access to the potential benefits derived from the intervention.

**Fig 1.**
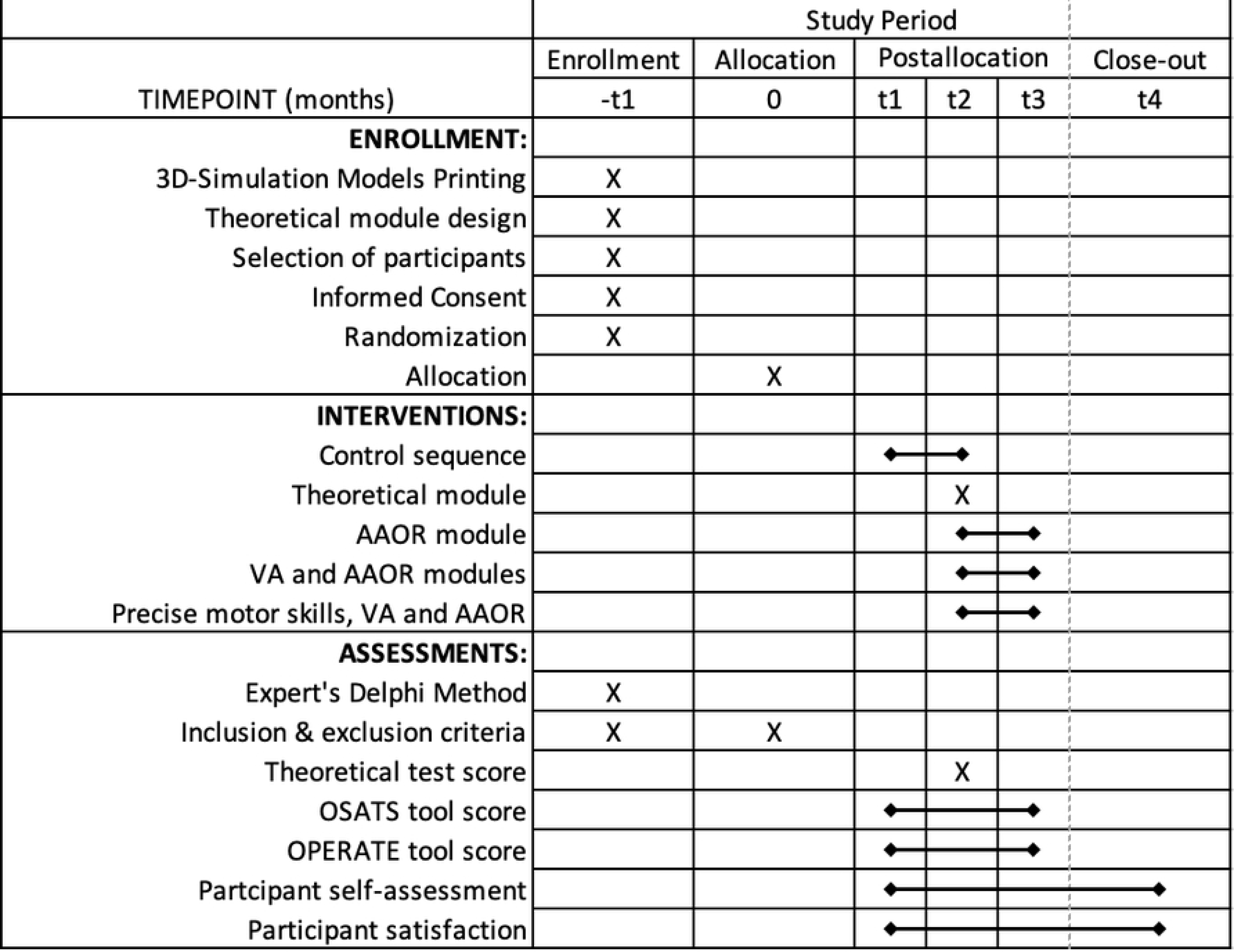
SPIRIT schedule of enrollment, interventions, and assessments.

**Fig 2.**
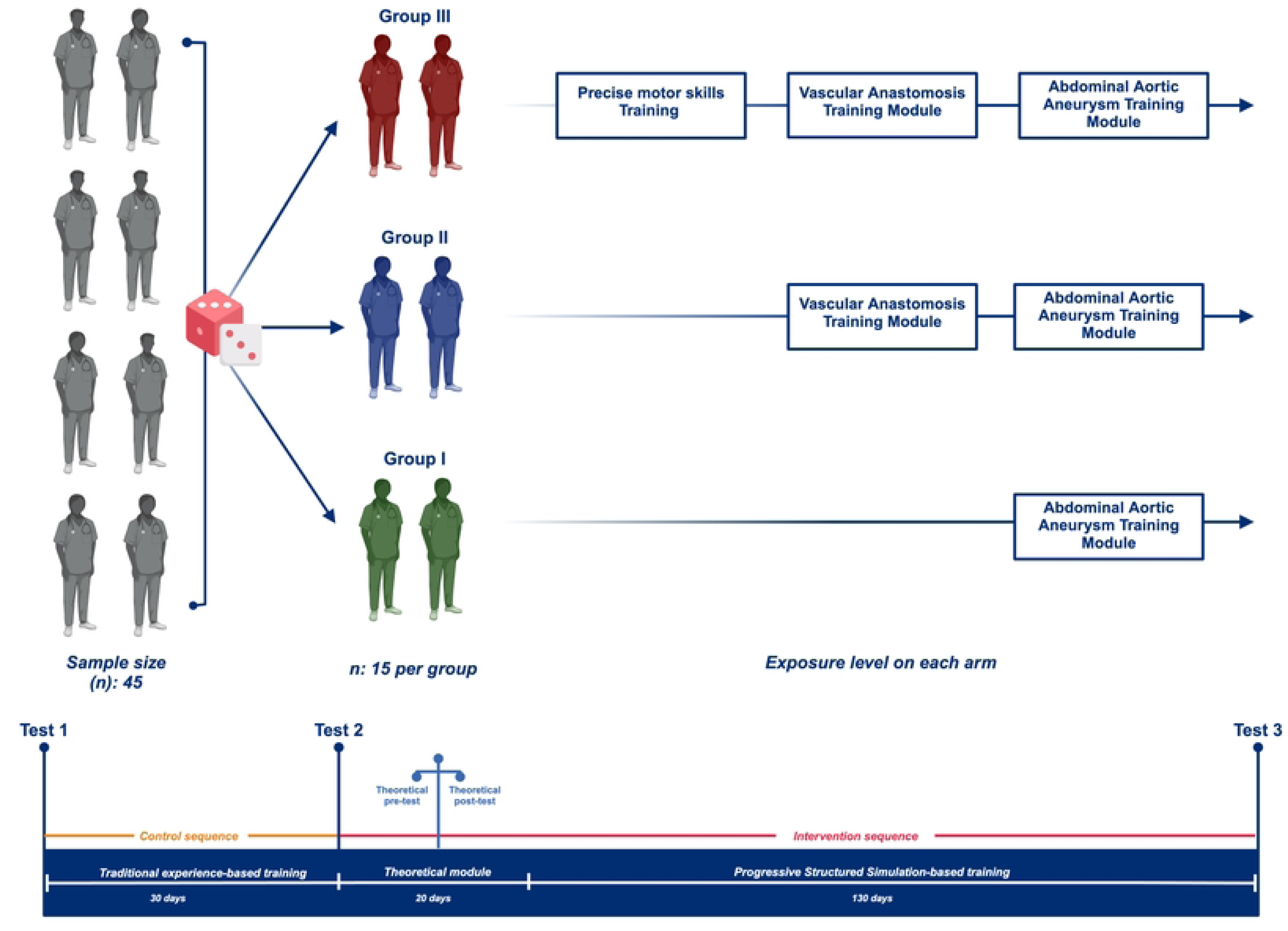
Graphic representation of the methodological design of the study.

In the control phase, participants will continue with experience-based traditional training and will not receive any simulation-based training intervention.

A theoretical module will be provided in the intervention phase to ensure the minimum required knowledge. The simulation-based training program in OVS consists of three courses: specific surgical skills, isolated surgical skills (VA), and a complex surgical procedure (AAOR). Participants will be randomized into 3 groups with different exposure to the simulation-based program. Group I will only receive the AAOR training module. Group II will receive the VA and AAOR training modules. Group III will receive precise motor skills, VA, and AAOR training modules (**Fig 2**.)

Each training module will consist of 2 practical sessions of simulation-based training per participant with a total of 6 hours. The sessions are organized according to the model established by Kopta et al. The first session is cognitive and integrative, where participants will observe an expert vascular surgeon performing the procedures, ask questions, and seek guidance. The remaining sessions will be autonomous (14).

### Sample Size Calculation

The minimum number of participants to achieve a significance estimate was made with the formula described by wang et al. n = [(Zα/2 + Zβ)/(d- δ))^2] [(p2(1-p2)/k + p1(1-p1)], were n represents the sample size per group (intervention and control)(15). Zα/2 = 1.28 is the critical z-value for the desired significance level (e.g., for a significance level of 10%). Zβ = 0.8 is the critical z-value for the desired power (e.g., for a power of 80%). δ = 16% margin of superiority. d = p1-p2 is the difference between the actual response rates of the intervention group. A superiority of 60% against 40% is expected. k = 1, same size between groups. Subsequently, we substitute into the formula that at least 39 participants (13 per group) would be needed. Additionally, 10% of enrolled participants are expected to drop out due to any circumstances that may arise and, therefore, not be analyzable. Thus, approximately 45 participants (15 per group) are needed.

### Participant selection

Participants will be selected from residents in general, vascular, and cardiothoracic surgery, considered related specialties that potentially benefit from the intervention (16,17). Participants must be in good health and free from conditions that could interfere with study participation. Severe illnesses, known allergies to intervention components, or medical conditions that hinder study compliance may lead to exclusion. Individuals unable to provide informed consent due to cognitive or legal incapacity will also be excluded. Additionally, those with prior experience in structured open vascular surgery simulations will be excluded to avoid bias from pre-existing familiarity.

The medical education department at Fundación Cardioinfantil-laCardio will disseminate the study through institutional emails to partner schools. Additionally, the study will be promoted on social media platforms such as LinkedIn, Instagram, and X. Potential participants will receive information about the study, its overall design, objectives, and expected benefits. Emphasis will be placed on the voluntary nature of participation. Only those who express a willingness to participate will be considered and provided with further information on the next steps. The process for expressing interest includes: (1) Completing an interest survey; (2) Receiving an email with more information, including informed consent, confidentiality agreement, and personal data consent form; (3) Responding to the email to confirm interest and submitting signed documents; and (4) Scheduling for randomization and a pre-test.

Participants will not be excluded under any circumstances based on sex, race, socioeconomic status, sexual orientation, university offering the residency program, or current level of training in medical residency. The invitation will not be sent or designed to include the name of a professor or person of authority so that communication is implemented neutrally and there is no influence from authority figures in the recruitment process. The disclosure process will be done through the official communication channels of the collaborating institutions involved in this study.

### Participant retention

When possible, program directors of the enrolled participants will be contacted to help reserve time frames. A schedule with the required time frames will be sent out from the beginning so that participants can adjust accordingly. A logistics team will frequently contact the participants to remind them of their session times, depending on the group they are allocated. “Make-up” days will be available for participants who are unable to attend a session, allowing them to complete it at an alternative time frame.

### Informed consent

Each participant will be asked to give written informed consent before participating in the study.

Researchers will provide detailed information about the study’s objectives, procedures, potential risks, and benefits. Participation will be emphasized as voluntary, with the freedom to withdraw at any time without negative consequences (**S2 Appendix)**.

### Randomization

Participants will be randomly allocated into one of the three groups using stratified randomization.

Participants will be stratified based on two factors: their residency level (from first to fourth year) and their residency type (General vs. Vascular or Cardiothoracic surgery). Initially, participants will be categorized into subgroups according to their year of residency. Within each residency level subgroup, participants will further be divided into their type of residency. After establishing these strata, participants within each stratum will be randomly assigned to one of the three study arms. This approach ensures a balanced representation of residency levels and types across all groups, enhancing the reliability and validity of the trial results.

The allocation sequence will be implemented using sealed envelopes and the randomization will be conducted in a single session after all participants have been enrolled. The sequence will be computer-generated using the application randomization.com. Each participant will receive their assignment through sequentially numbered, opaque, sealed envelopes, ensuring that the allocation sequence remains concealed until the interventions are assigned.

### Blinding

Evaluators responsible for assessing the efficacy of the different interventions will undergo a single blinding procedure. These evaluators will be external and independent from the researchers and unaware of the participants’ group assignments. Privacy regarding participants’ group assignments will be maintained through coding and presentation of de-identified data. Data will be presented as a videotape without voices, with glove usage, and image distortion to prevent the identification of personal details such as gender, race, or other characteristics. Under no circumstances will the identity of the participants be revealed to the evaluators.

### Simulation models

A 3D printed AAOR simulation model is being developed (**Fig 3 A and B**). We aim to create 3D printed models from AngioCT images of three patients with unruptured AAA requiring AAOR at our institution.

**Fig 3.**
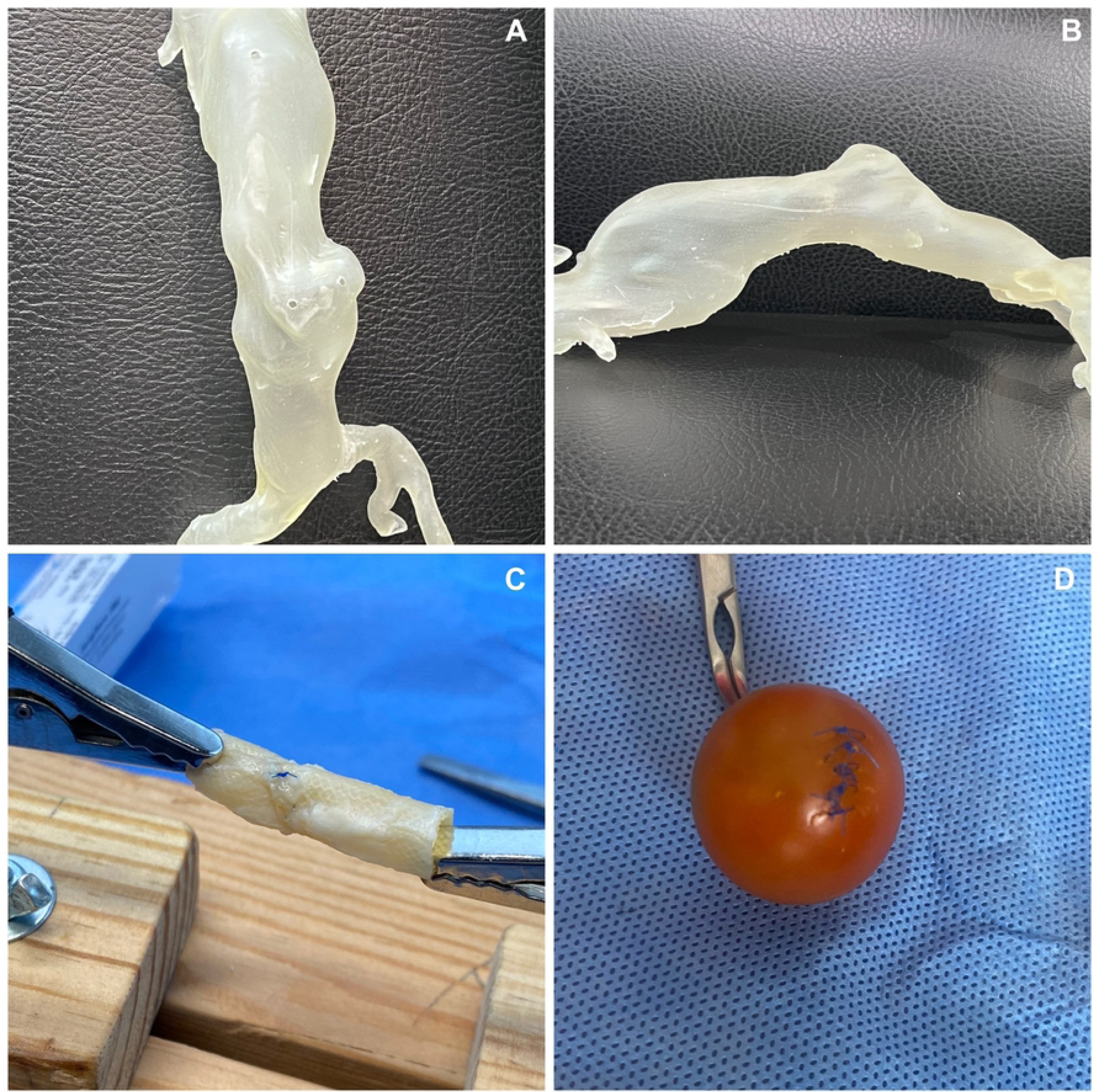
Preliminary simulation models. (A-B) AngioCT 3D-printed models of a real-life patient with unruptured AAA (C) VA models using wooden platforms suspended by clamps at adjustable distances. (D) Specific micro-surgical skills simulation models will incorporate tools like elongated cherry tomatoes, prolene 6.0, and basic surgical instruments.

Before using the diagnostic images, patients will be informed of the purpose of the study, ensuring that it does not affect their medical or surgical treatment. Patients must understand, accept, and sign an informed consent form to use their images in the creation of 3D models for research use only and anonymize the metadata of the DICOM files.

The most suitable AngioCT images will be selected based on criteria such as image quality (including slice thickness, number, and spacing of slices) and the patient’s clinical case, focusing on an unruptured AAA. This collaborative effort involves vascular surgeons and engineers using specialized software, Mimics Innovation Suite 25.0 (Materialise, Belgium), for medical image processing.

Following image selection, the infrarenal abdominal aorta segmentation employs semi-automated threshold-based methods to isolate the blood volume. A 3D reconstruction is conducted, and the resulting file is exported in STL format. Subsequently, the aortic wall thickness is determined, set between 1.25 - 2.1 mm, based on the population’s average abdominal aortic wall thickness to ensure a realistic feel for suturing in the 3D printed mode (18,19). It is assumed that the wall thickness throughout the model will be uniform.

The 3D reconstruction model of the abdominal aorta will be divided into three segments: 1) the region of the aneurysm, 2) the region above the aneurysm, and 3) the region below the aneurysm. This division is intended for 3D printing of the model. Segments 2 and 3 (the regions above and below the aneurysm) will be printed in rigid material, while segment 1 (the aneurysm region) will be printed in flexible and elastic material. This approach optimizes resources since the aneurysm region will constantly change during practice. For 3D printing of segment 1 (the aneurysm region), PolyJet and LCD technology with Agilus30 (Stratasys, Ltd., USA) and flexible UV170TR (Photocentric, UK) photopolymer resins will be used, respectively, to obtain models with elastic and flexible material to allow suturing.

For segments 2 and 3, PolyJet technology with rigid VeroBlue material (Stratasys, Ltd., MN, USA) will be used (**Fig 4**).

**Fig 4.**
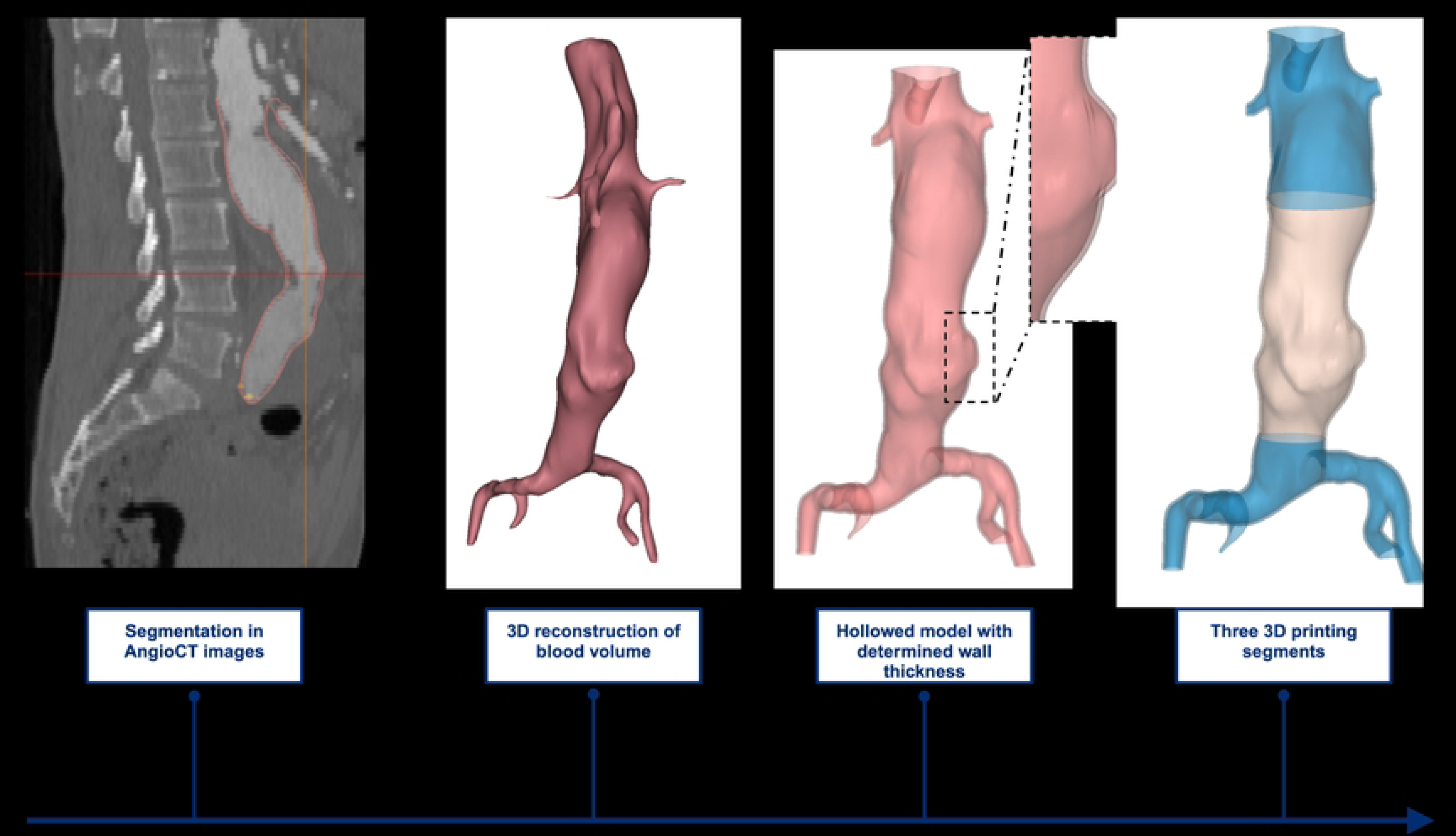
Graphical representation of the models’ selection process, reconstruction, and 3D segmentation.

VA simulation models will be constructed using wooden platforms suspended by clamps at adjustable distances. This approach requires minimal material for the VA (**Fig 3C**).

Specific micro-surgical skills simulation models will require tools facilitating the development of expected competencies, including elongated cherry tomatoes, prolene 6.0, and basic surgical instruments. (**Fig 3D**).

### Evaluation instruments

A range of tools previously validated for assessing surgical skills in OVS will be utilized.

These tools include standardized grading rubrics and will involve external evaluators.

The OPEn aortic aneurysm Repair Assessment of Technical Expertise (OPERATE) tool, developed and validated by Nayahangan et al., is a structured assessment instrument designed to ensure competency in AAOR (11,20). The assessment is divided into distinct domains: patient positioning, incision and exposure, proximal and distal control, aneurysm sac preparation, graft implantation, and closure. Each domain consists of tasks rated on a numerical scale (1-3-5), with higher scores indicating better performance. The validation demonstrated that the tool effectively distinguishes between varying levels of expertise, making it a reliable and robust method for assessing surgical competence in the AAOR (11).

The Objective Structured Assessment of Technical Skills (OSATS) tool is widely recognized for evaluating surgical trainees’ technical skills, including those in the vascular surgery (21,22). Each task is rated using a numerical scale and task-specific checklists, ensuring a comprehensive evaluation. According to Lladó Grove et al., the OSATS tool includes criteria such as respect for tissue, time and motion efficiency, instrument handling, procedure knowledge, operation flow, and use of assistants (21). These criteria are rated on a Likert scale, typically 1, 3, or 5, where higher scores indicate better performance. Sidhu et al. further expanded on the tool’s utility by integrating it into a comprehensive vascular skills assessment, demonstrating its effectiveness in distinguishing between different levels of trainee expertise (22).

Also, a flow simulator for anastomosis quality assessment will mimic the rheological characteristics of the abdominal aorta surgery. Its purpose is to evaluate the quality of vascular anastomosis via an anastomotic leakage test employed in both introductory surgical and complex surgical procedure courses. This simulator comprises flow measurements, collection, and pressure measurement devices, ensuring precise hydrostatic pressure measurement (+/—0.3 kPa). The saline solution will serve as the medium.

### Data collection instrument

Data for the study will be collected and securely stored online using the REDCap web application. Only anonymized data, identified by randomization labels, will be uploaded to the database to ensure privacy. The primary data sources include participant surveys, external assessors’ evaluations, and logistics personnel’s objective data. No secondary data will be collected.

### Outcomes

Each participant will be the object of observation and measurement for this study. We adhere to Messick’s validation framework (7), so the evaluation scores must correlate with known measures of competence, and pass/fail grades will be established. The assessment of the intervention’s effects will depend on the intervention arranged in each study phase.

For the theoretical module, a traditional evaluation through multiple-choice questions will be used (24). A passing score of 3.0 out of 5.0 points will be utilized, which means a better outcome. Two theoretical tests will assess the efficacy of this intervention before and after its application.

For the specific micro-surgical skills and isolated surgical skills (VA), the “OSATS tool” will be used (7,21,22). We adhere to the passing score established by Chipman et al. of 75% of the “minimum required” skill according to each module (23). A higher score means a better outcome.

For the complex surgical procedure (AAOR), the “OPERATE tool” will be used (11). We adhere to the established passing score of 27.7 points. A higher score means a better outcome.

Standardizing minimum and maximum volumes will also establish thresholds for acceptable leakage levels for the anastomosis quality with the flow simulator.

In the control phase, two tests will be conducted to collect data on their performance and technical surgical skills without the direct influence of the educational intervention. For the intervention phase, the second test of the control phase will serve as the starting baseline, and a third test will be conducted at the end of the simulation-based surgical training.

Before starting each session/test in all phases, structured surveys will be completed to establish self-assessment, satisfaction, and confounding factors such as exposure to vascular surgical procedures (intervention). A Likert scale with a minimum value of 1 and a maximum value of 10 will be used. A higher score means a better outcome.

### Data management plan

Data will be managed in compliance with Colombian law 1581 of 2012, ensuring confidentiality and anonymization. The REDCap platform will secure data, with access restricted to authorized users. Periodic audits (monthly) will maintain data quality and address discrepancies. Precise and standardized instructions will be provided to all participants before each course and simulation test, verified during the pilot test. Robust logistical procedures will manage the pilot test, identifying issues before implementation. Data collection will follow a standardized methodology, ensuring secure and consistent entry. The research center at Fundación Cardioinfantil – La Cardio will exclusively protect and store data. Study documents will be archived or stored on institutional servers for five years post-study, then transferred to long-term storage for up to 15 years before destruction.

### Statistical analysis

Both conventional and Machine Learning methods will be utilized. The analysis will begin with exploratory data analysis, including calculating descriptive statistics such as mean, median, and standard deviation for the self-assessment and satisfaction surveys, controlling confounding factors, and characterization of participants. Correlation analysis (Pearson or Spearman, as appropriate) will be performed to explore relationships between participant characteristics and skill acquisition, retention, and satisfaction. Visualizations, such as histograms and bar charts, will be created to represent the distribution of these results and frequency analysis will be performed to evaluate the prevalence of various factors and characteristics within the intervention groups. Anomaly detection techniques like Isolation Forest or One-Class SVM will be used to detect unusual performance or deviations from expected outcomes.

For the primary objective, a comparative analysis will be conducted between the three randomized groups. A repeated-measures ANOVA will be used to analyze differences in skill acquisition within and between groups. Post-hoc pairwise comparisons will be conducted to identify specific group differences. A significance level of p < 0.05 will be set for all analyses.

To assess technical surgical skills in OVS before and after the implementation of the structured training program paired t-tests will be used to compare pre-and post-intervention skill levels within each group. To analyze differences in skill acquisition attributed to the varying levels of simulation exposure among different groups within the program a subgroup analysis will be performed to evaluate the impact of different simulation exposure levels on skill acquisition, ANCOVA will be used to control for potential confounders such as baseline skill level, year of residency, and previous surgical experience. To evaluate the reaction and satisfaction of residents with the structured, progressive simulation-based training program in OVS differences in satisfaction levels between groups will be explored using Chi-square tests or Kruskal-Wallis tests, depending on data distribution.

Additionally, predictive modeling techniques such as regression trees and random forests will be employed to assess and forecast skill acquisition efficacy based on participants’ demographics, prior experience, and exposure to various training modules. Clustering analysis methods, including k-means clustering and hierarchical clustering, will segment participants into distinct groups based on their skill acquisition patterns and responses to the training interventions. This will help identify subgroups that exhibit similar responses to different training approaches. Dimensionality reduction techniques, such as Principal Component Analysis (PCA) and t-SNE, will simplify the data complexity and enhance the visualization of relationships between training exposure, skill acquisition, and other variables, thereby highlighting the key factors contributing to successful skill development.

### Analysis population and missing data

The analysis population will be based on the intention-to-treat (ITT) principle, meaning all participants will be included in the analysis according to their original randomization group, regardless of their retention or adherence to the protocol. This approach aims to maintain the randomization integrity and reflect real-world effectiveness. Missing data, primarily due to participant dropouts and loss of retention, will be addressed with a combination of methods. While imputation techniques will be predominantly employed to estimate missing values, ensuring that the analysis utilizes the maximum amount of available data, a sensitivity analysis will also be conducted to assess how different methods for handling missing data impact the study results. This comprehensive approach will help to evaluate the robustness of the findings and mitigate potential biases introduced by incomplete data.

### Safety considerations

The primary risk of this study arises from the hierarchical setting, and we recognize the importance of clarifying this aspect. Although Fundación Cardioinfantil – La Cardio professors will facilitate the cognitive and integrative simulation sessions, it is crucial to emphasize that independent external evaluators will conduct performance evaluations in a fully anonymous and standardized manner. This external evaluation will not affect the participants’ performance or status within their respective training programs, ensuring impartiality in the assessment.

Handling surgical instruments during simulation sessions may raise concerns about potential personal injuries. To minimize this risk, rigorous safety protocols that have been standardized and established by the advanced simulation center at Universidad del Rosario will be followed. Detailed guidance on the safe handling of instruments will be provided during the sessions, and expert professionals will ensure constant supervision. Additionally, emergency measures and procedures are in place to address any unexpected situations and ensure participants’ physical safety.

### Delphi method

Following Messick’s framework of validity, a systematic type 2 Delphi method will be used before starting the clinical trial or pilot test (24). A heterogeneous group of anonymous expert vascular surgeons will evaluate a standardized questionnaire, adapted for multiple rounds, to select content components for the theoretical and practical modules. Each round will refine the questionnaire based on expert feedback until consensus is achieved.

Consensus will be defined as agreement by 80% of the experts (4:1 ratio), and lack of consensus will be defined as less than 20% agreement (4:1 ratio). A maximum of three rounds will be conducted (initial round plus two extensions). It will be eliminated if a question fails to achieve more than 80% agreement. The final round will conclude the Delphi process if consensus is reached, or a live meeting will be held if needed.

### Pilot study

Before fully implementing the structured progressive simulation-based open vascular surgery training program, a pilot test will be conducted with 6 participants to identify and address potential logistical and methodological issues. Objectives include standardizing simulation parameters, evaluating protocol logistics, and training external evaluators. The pilot study will follow the same stages as the full protocol but over a shorter period. Key focus areas include participant selection, randomization, blinding effectiveness, 3D model printing accuracy, anastomosis quality measurement, theoretical virtual module comprehension, technological support, simulation training logistics, and data collection on REDCap. This phase aims to ensure the quality and reliability of the entire study by resolving any identified problems.

### Status and timeline

Recruitment has yet to begin; we expect to finish recruitment on 31 September 2024. The control phase will probably begin in October 2024. The intervention phase may run from November 2024 to March 2025. The analysis and scientific writing may span from March to June 2025.

### Trial registration

This study protocol was registered with the NCT-ID: NCT06452901 on www.ClinicalTrials.gov using the account of the research center at Fundación Cardioinfantil – laCardio.

### Data availability

Due to participants’ privacy and Colombian law concerns, the data will not be available for public access but may be available from the corresponding author upon reasonable request.

## Discussion

Vascular surgery has become a highly complex specialty requiring more competent surgeons (3–5). Despite this, current surgical training programs are often criticized for their limited ability to impart technical skills in open vascular surgery, mainly due to varying clinical case exposures for each trainee (7). Simulation strategies have proven effective in enhancing surgery’s technical and theoretical learning curve, especially in the early stages of training (16,17). However, existing evidence on simulation-based programs for OVS is mainly limited to non-experimental, heterogeneous observational studies conducted with North American and European trainees (7).

Implementing a micro-curriculum for a progressively structured simulation-based training program in OVS can promote the acquisition of critical surgical competencies in future vascular surgeons. Additionally, it seeks to standardize the learning process for surgical skills in OVS, establishing quality benchmarks for surgical programs involving these procedures. In the medium to long term, this methodology could encourage adopting and adapting this strategy in medical centers and universities, both nationally and internationally, within their surgical training curricula. This standardization ensures that all trainees receive a consistent and high-quality educational experience, regardless of their training location or exposure to OVS (7).

Developing reusable simulation models for AAOR represents a significant innovation in applying 3D printing technology in vascular surgery. Traditionally, 3D printing has been primarily used for planning and training in endovascular techniques (25). However, our study may demonstrate that this technology can also significantly enhance training in OVS. By creating accurate and reusable simulation models, we can provide an efficient tool for AAOR training, offering a precise and personalized representation of patient anatomy and facilitating the planning and execution of AAOR procedures.

Our study’s sequential structure aligns with Kirkpatrick’s levels 1 and 2 of training evaluation efficacy, focusing on immediate learner reactions and the extent of learning (7). This approach is crucial for assessing the immediate impact of the training program on the participant’s skills and knowledge. Moreover, it provides a basis for future studies to evaluate the transfer of skills to the operating room (Kirkpatrick level 3) and subsequent surgical outcomes in patients (Kirkpatrick level 4) (7). By enhancing surgical skills through a structured simulation-based program, we anticipate improved surgical outcomes, reduced complications, and enhanced patient safety.

The three-arm design of this study ensures that all participants have access to the potential benefits derived from the intervention, thereby maximizing the educational value for each participant. This approach is particularly advantageous as it aligns with current evidence supporting the benefits of medical simulation in the training process of surgical residents (16,17). This structure provides a robust framework for evaluating the differential impacts of the intervention but also ensures that every participant is provided with potentially valuable training opportunities.

Also, following the Messick validation framework is critical in ensuring the robustness and credibility of simulation- based training programs, particularly in the context of OVS (7). According to Lawaetz et al., this framework provides a comprehensive approach to validating educational interventions (7). By adhering to it, our study aims to ensure that OVS’s structured, progressive simulation-based training program is rigorously evaluated for its efficacy, fairness, and applicability. This approach enhances the reliability of the findings and supports the standardization of surgical training.

Given the design and nature of this study, multiple biases may arise, necessitating effective identification and control strategies. Learning bias is a significant concern, with participants potentially experiencing period effects caused by changes in participants’ conditions, such as varying exposure to surgical practices or self-learning, which will be controlled through pre-session surveys to identify confounding factors and ensure adequate randomization among the intervention groups. Sequence effects, influenced by the order of study periods, will be managed by adopting a logical phase based on the difficulty of “unlearning” acquired knowledge, thus providing a natural progression for the learning process.

Integrating cognitive and technical lessons through a blended learning methodology is essential in modern surgical education. Jayakumar et al. highlight the significant benefits of e-learning in surgical education, noting that it allows for flexible, self-paced learning while providing access to a wide range of resources and interactive content (26). Additionally, Malas et al. emphasize the importance of visualization in simulation training for VA (14). Their findings indicate that visual learning aids, such as detailed anatomical models and procedural simulations, significantly improve trainees’ technical skills. Combining cognitive lessons delivered through blended learning with hands-on technical training in a simulated environment creates a comprehensive educational experience that enhances understanding and practical proficiency.

Selection bias, which may occur if participant assignment to groups is not genuinely random, will be mitigated by stratified randomization by ensuring that key variables, such as baseline characteristics or covariates, are evenly distributed across different treatment groups. Consistent application of inclusion and exclusion criteria throughout the study will further reduce this bias. The selection of participants within these specialties is crucial to ensure that the study subjects have sufficient medical knowledge and skills to evaluate the efficacy of the proposed educational interventions (27). Additionally, including residents from all years of training helps to further reduce selection bias by ensuring a diverse range of experience levels, which can enhance the generalizability of the findings. Importantly, each participant will serve as their control, thus minimizing the potential for confounding factors and ensuring that individual variations in baseline knowledge and skills do not affect the overall outcomes.

The data audit team will minimize labeling bias, which results from systematic errors in participant classification, by carefully reviewing and verifying labels. Attrition bias, which arises if dropouts differ systematically from remaining participants, will be addressed by calculating additional sample size to ensure sufficient participant numbers at the study’s conclusion. Measurement bias will be controlled using blinded evaluators and standardized evaluation instruments, with thorough training provided to ensure consistency and accuracy.

Despite the meticulous design, several limitations must be considered when interpreting the results. One significant limitation is the inability to blind researchers and participants who conduct and supervise the simulation courses, potentially introducing unconscious biases. The sequential and educational nature of the study presents a confounding variable, as it is impossible to “unlearn” previously acquired knowledge. Additionally, the study is designed to measure short-term learning impacts (Kirkpatrick levels 1 and 2) but does not assess longer-term outcomes (Kirkpatrick levels 3 and 4), which involve patient interactions and follow-up assessments (7). Financial considerations also pose a limitation, as acquiring high-quality simulators and implementing courses can incur substantial costs, potentially affecting the program’s scalability in budget-constrained settings. Various funding avenues and cost-reduction strategies are proposed to address these challenges. However, the protocol has been designed to address these concerns effectively, ensuring the validity and reliability of the findings.

We are committed to registering and publishing this study protocol as an essential practice that promotes transparency, reproducibility, and control of publication bias. This practice ensures that studies with significant and non-significant results are acknowledged, providing a comprehensive and unbiased view of the evidence. Furthermore, accessible registration facilitates detailed methodological information sharing, essential for peer review, replication, and meta-analyses. This openness promotes collaboration, prevents unnecessary duplication, and enhances trust in future publications in indexed journals and presentations at medical conferences of our work (28).

## List of abbreviations

AAA: Abdominal Aortic Aneurysms, EVAR: Endovascular Aneurysm Repair, AAOR: Abdominal Aortic Aneurysms Open Repair, OVS: Open Vascular Surgery, VA: Vascular Anastomoses, OPERATE: OPEn aortic aneurysm Repair Assessment of Technical Expertise, OSATS: Objective Structured Assessment of Technical Skills.

## Declarations

### Version identifier

Version 4.0, emission date 14/08/2024.

### Ethics approval and consent to participate

This study will be conducted under ethical norms and adhere to Resolution 8430 of 1993 of the Ministry of Health of the Republic of Colombia and Law 1581 of 2012 regarding data protection. It was classified as minimum-risk research. The Institutional Clinical Research Ethical Committee of Fundación Cardioinfantil-La Cardio approved this study protocol (Ref Number: 047-2023). Informed consent will be obtained from all participants enrolled in the study (CEIC-0530-2026).

### Presentation information

A preliminary version of this protocol was presented at the 2024 Scientific Sessions Vascular Discovery: From Genes to Medicine of the American Heart Association (AHA) poster session and was awarded the Paul Dudley White International Award.

### Authors’ contributions

AVS, JCM, COM, JGBC, and SGG conceived the idea. AVS, CAPS, JVAM, SUR, and JTP wrote the manuscript. AVS, MMPJR, CJPR, COM, and PACR planned the methodology. DLCR and COM planned the methodological analysis. YFBM, CEPC, and JCB designed the simulators. AVS, JAVM, ECB, MMPJR, SGG, JCM, and JGBC planned the program content. SGG, JCM, JGBC, JCB, MMPJR, YFBM, and COM critically revised the manuscript. All authors read and approved the manuscript. We adhere to the authorship criteria of the International Committee of Medical Journal Editors (ICMJE).

## Acknowledgments

We would like to acknowledge the help and work from César Camilo Medina Suárez MD, Julian M. Corso-Ramírez MD., Euzcady Rafael López Martínez MD., María A. Rodríguez-Soto MSc, PhD., Javier Navarro Rueda MSc, PhD., Camila Irene Castro Páez MSc, PhD.

